# Preliminary evidence from a multicenter prospective observational study of the safety and efficacy of chloroquine for the treatment of COVID-19

**DOI:** 10.1101/2020.04.26.20081059

**Authors:** Mingxing Huang, Man Li, Fei Xiao, Jiabi Liang, Pengfei Pang, Tiantian Tang, Shaoxuan Liu, Binghui Chen, Jingxian Shu, Yingying You, Yang Li, Meiwen Tang, Jianhui Zhou, Guanmin Jiang, Jingfen Xiang, Wenxin Hong, Songmei He, Zhaoqin Wang, Jianhua Feng, Changqing Lin, Yinong Ye, Zhilong Wu, Yaocai Li, Bei Zhong, Ruilin Sun, Zhongsi Hong, Jing Liu, Huili Chen, Xiaohua Wang, Zhonghe Li, Duanqing Pei, Lin Tian, Jinyu Xia, Shanping Jiang, Nanshan Zhong, Hong Shan

## Abstract

**Background:** Effective therapies are urgently needed for the SARS-CoV-2 pandemic. Chloroquine has been proved to have antiviral effect against coronavirus in vitro. In this study, we aimed to assess the efficacy and safety of chloroquine with different doses in COVID-19.

**Method:** In this multicenter prospective observational study, we enrolled patients older than 18 years old with confirmed SARS-CoV-2 infection excluding critical cases from 12 hospitals in Guangdong and Hubei Provinces. Eligible patients received chloroquine phosphate 500mg, orally, once (half dose) or twice (full dose) daily. Patients treated with non-chloroquine therapy were included as historical controls. The primary endpoint is the time to undetectable viral RNA. Secondary outcomes include the proportion of patients with undetectable viral RNA by day 10 and 14, hospitalization time, duration of fever, and adverse events.

**Results:** A total of 197 patients completed chloroquine treatment, and 176 patients were included as historical controls. The median time to achieve an undetectable viral RNA was shorter in chloroquine than in non-chloroquine (absolute difference in medians −6.0 days; 95% CI −6.0 to −4.0). The duration of fever is shorter in chloroquine (geometric mean ratio 0.6; 95% CI 0.5 to 0.8). No serious adverse events were observed in the chloroquine group. Patients treated with half dose experienced lower rate of adverse events than with full dose.

**Conclusions:** Although randomised trials are needed for further evaluation, this study provides evidence for safety and efficacy of chloroquine in COVID-19 and suggests that chloroquine can be a cost-effective therapy for combating 102 the COVID-19 pandemic.

## Introduction

The coronavirus disease 2019 (COVID-19) emerged in late 2019, originating from Wuhan China^1,2^. The responsible virus, severe acute respiratory syndrome coronavirus 2 (SARS-CoV-2), belongs to a distinct clade from the human severe acute respiratory syndrome CoV (SARS-CoV) and Middle East respiratory syndrome CoV (MERS-CoV)^3^. It has become a global pandemic, affecting over 100 countries with more than 240,000 confirmed cases and over 10,000 deaths globally as of March 20, 2020, calling for an urgent demand of effective treatment.

Chloroquine has been proved effective in vitro to inhibit the replication of SARS-CoV^4^, HCoV-229E^5^, and the newly discovered SARS-CoV-2^6,7^. To evaluate the efficacy and safety of chloroquine for COVID-19, we previously conducted a single-arm pilot clinical study with 10 patients (Huang et al. Journal of Molecular Cell Biology, in press). Encouragingly, all patients achieved undetectable level of viral RNA within 14 days without serious adverse events. These results led us to conduct a multicenter prospective observational study in adult patients with COVID-19 to assess the efficacy and safety of chloroquine for COVID-19.

## Result

### Patients

Of the 233 enrolled patients for chloroquine, 197 (84.5%) completed treatment and were included in the final analysis (**Figure 1, study flowchart; Supplementary Table 1**). Of the 182 patients collected as historical controls, 176 (96.7%) were included in the final analysis. Their baseline demographic and clinical features are listed in **Table 1**. The median age of patients were 43 years (inter-quartile range [IQR], 33 to 55 years) in the chloroquine group and 47.5 years (IQR, 35.8 to 56 years) in the non-chloroquine group. Across the two treatment groups, the majority patients were classified as moderate cases (93.4% in chloroquine; 89.2% in non-chloroquine)^8^. Chloroquine was added into China’s Diagnosis and Treatment Guidelines of COVID-19 later than the other therapies used in the non-chloroquine group. Therefore, we observed longer interval time between symptom onset and treatment initiation in chloroquine versus non-chloroquine (absolute difference 4 days; 95% CI 2 to 6 days; *P* < 0.0001). In addition, due to the rapid rise of patients in Wuhan and established mobile hospital in early February, the interval time between symptom onset and treatment initiation in Wuhan (median 17 days, IQR 10.5 to 21 days) is longer than that in Guangdong Province (median 5 days, IQR 3 to 10 days; **Table 1**). In the subgroup of patients from the Fifth Affiliated Hospital of Sun Yat-sen University (SYSU5), we obtained and evaluated the viral load at baseline between chloroquine (N=21) and non-chloroquine (N=8) group and did not observe statistically significant difference (absolute difference in medians = 2.93, 95% CI −0.8 to 6.6, p = 0.09).

**Table 1.** Baseline characteristics in chloroquine and non-chloroquine among people with COVID-19.

|  | **chloroquine**<br>**(N=197)** | **Non-chloroquine**<br>**(N=176)** |
| --- | --- | --- |
| Guangdong, N (%) | 118 (60) | 96 (54) |
| Hubei, N (%) | 79 (40) | 80 (46) |
| Age, mean (SD) | 43.8 (13.1) | 45.6 (13.5) |
| Age ≤ 65 | 190 (96) | 171 (97) |
| Age > 65 | 7 (4) | 5 (3) |
| Female sex, N (%) | 101 (51) | 97 (55) |
| Clinical manifestation<sup>†</sup>, N (%) |  |  |
| Mild | 9 (5) | 5 (3) |
| Moderate | 184 (93) | 157 (89) |
| Severe | 4 (2) | 14 (8) |
| Comorbidities, N (%)<sup>*</sup> |  |  |
| Hypertension | 13 (17) | 11 (17) |
| Type 2 diabetes | 4 (5) | 5 (8) |
| Interval time from symptom onset to treatment initiation, median (IQR) |  |  |
| Guangdong | 7 (3, 10.8) | 4 (2, 7) |
| Hubei | 19 (17, 24.5) | 11 (7, 16) |
| Body temperature, median (IQR), °C | 36.7 (36.5, 37.0) | 36.6 (36.4, 37.3) |
| Pneumonia from chest CT, N (%)<sup>§</sup> | 173 (89) | 137 (93) |

<sup>\*</sup> The number of patients with valid record of comorbidities are 78 in chloroquine group and 66 in non-chloroquine group
<sup>§</sup> The number of patients with valid record of chest CT image are 194 in chloroquine group and 148 in non-chloroquine group.
<sup>†</sup> clinical manifestation type definitions: 1) Mild, mild clinical symptoms with no signs of pneumonia on chest radiological imaging; 2) Moderate, fever, respiratory symptoms, imaging with pneumonia changes; 3) Severe, meet any of the following criteria: shortness of breath, respiratory rate > 30 times per minute, resting stable oxygen saturation in fingertip < 93%, oxygenation index < 300, pulmonary imaging showed that the lesion progressed significantly more than 50% within 24-48 hours; 4) Critical, if any of the following occurs: respiratory failure requiring mechanical ventilation; shock, concurrent with other organ failure requires intensive care.

**Figure 1.**
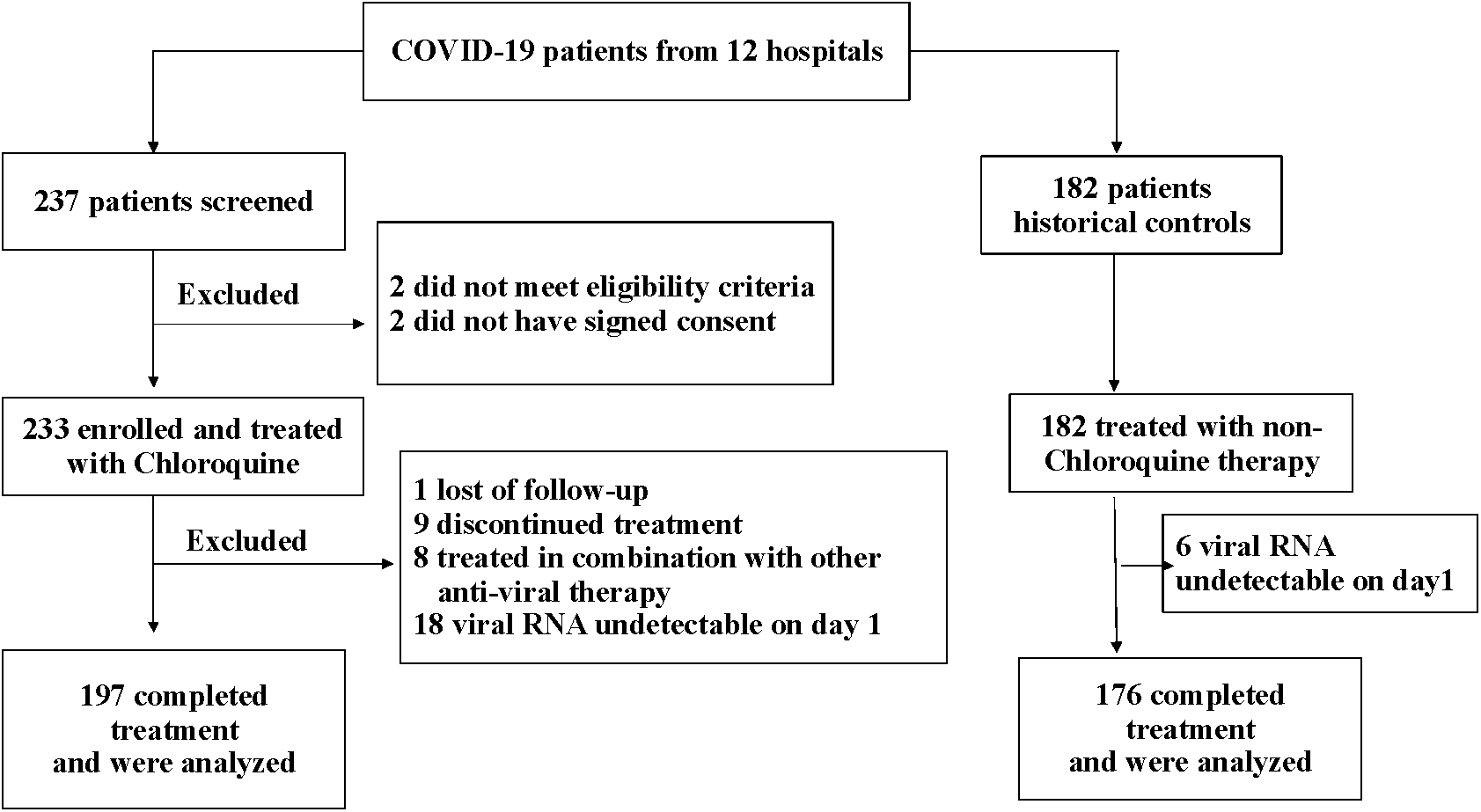
Study flowchart.

### Outcomes

In the analysis of the full study population, patients in the chloroquine group have an accelerated time to undetectable viral RNA from that of patients in the non-chloroquine group (absolute difference in medians −5.4 days; 95% CI −6 to −4; *P* < 0.0001; **Figure 2**). Secondly, by day 10 and day 14 since treatment initiation, higher proportion of patients had undetectable viral RNA in the chloroquine group (91.4% and 95.9% respectively; **Table 2**) comparing to the non-chloroquine group (57.4% and 79.6% respectively; **Table 2**). In the aspect of clinical manifestations, we found that the duration of fevers is shorter in chloroquine versus non-chloroquine among patients experienced fever symptom (geometric mean ratio 0.6; 95% CI 0.5 to 0.8; *P* = 0.0029; **Supplementary Figure S1**). To note, the antipyretic effects of chloroquine may have also contributed to this result. We observed no difference in the length of hospital stay (**Supplementary Figure S2**). No patient died or admitted to ICU either in the chloroquine group or in the non-chloroquine group. There are 1 patient in the chloroquine group experienced aggravated symptoms from moderate to severe, while 9 patients in the non-chloroquine group have the same aggravated experience. All of the 10 patients eventually were tested negative for the viral RNA within the study period.

**Table 2.** Outcomes in the overall population with confirmed SARS-CoV-2 infection^§^.

|  | **chloroquine**<br>**(N=197)** | **Non-**<br>**chloroquine**<br>**(N=176)** | **Difference**<br>**(95% CI)<sup>†</sup>** | **P value** |
| --- | --- | --- | --- | --- |
| Time to undetectable viral RNA, median no. of days (IQR) | 3.0<br>(3.0, 5.0) | 9.0<br>(6.0, 12.0) | -6.0<br>(-6.0, -4.0) | < 0.0001 |
| Patients with undetectable viral RNA by, N (%) |  |  |  |  |
| Day 10 | 180.0<br>(91.0) | 101.0<br>(57.0) | 34.0<br>(25.6, 42.9) | < 0.0001 |
| Day 14 | 189.0<br>(96.0) | 140.0<br>(80.0) | 16.0<br>(9.2, 23.3) | < 0.0001 |
| Duration of fever*, no. of days, geometric mean (CV) | 1.2<br>(53.5) | 1.9<br>(110.0) | 0.6<br>(0.5, 0.8) | 0.0029 |
| Length of hospital stay, median no. of days (IQR) | 19.0<br>(16.0, 23.0) | 20.0<br>(15.8, 24.0) | -1.0<br>(-3.0, 0.0) | 0.25 |

Abbreviations: CI, confidence interval; IQR, inter-quartile range; CV, coefficient of variation.
<sup>§</sup> Definitions of outcomes are listed in Supplemental Methods.
<sup>†</sup> 95% CI for continuous variables are calculated by bootstrapping. 95% CI for binary variables are calculated with Wilson method. The difference for duration of fever is geometric mean ratio of chloroquine group to non-chloroquine group. The differences for all other variables are the absolute difference between chloroquine group and non-chloroquine group.
\* The number of patients had at least one day of fever is 42 and 51 in the chloroquine and non-chloroquine group respectively.

**Figure 2.**
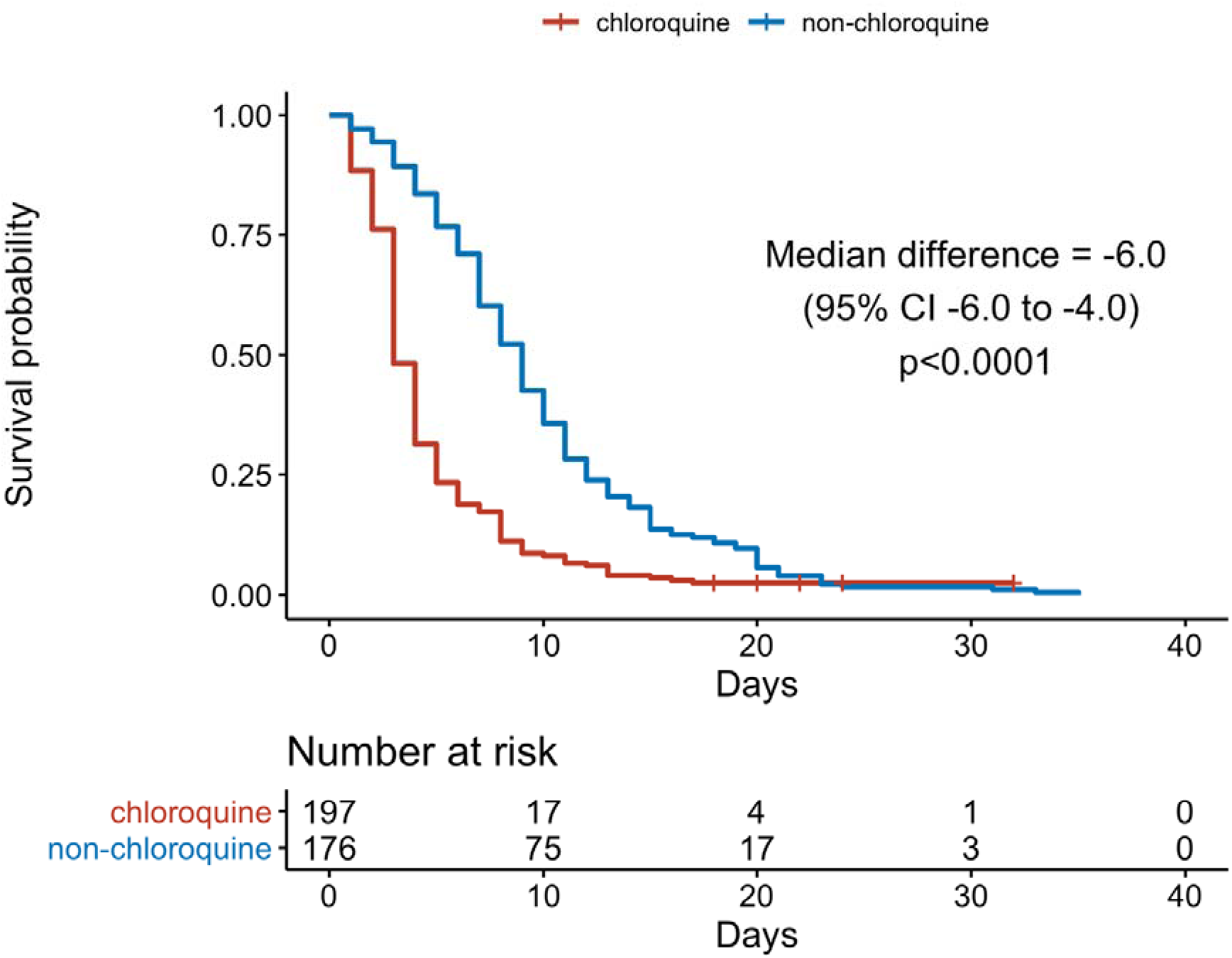
Kaplan-Meier curve for time to undetectable viral RNA comparing treatment groups.

Due to the significant difference observed in clinical classification between chloroquine and non-chloroquine group at baseline, we further analyzed the primary and secondary outcomes in patients with moderate symptoms only. The number of patients in mild or severe subgroup were too few to compare. The benefit of chloroquine in viral suppression is consistent with the full analysis, except for non-significant difference observed for the proportion of patients with undetectable viral RNA by day 14 (**Supplemental Table 2**).

In post hoc analysis, we examined the effect of chloroquine on the time to undetectable viral RNA stratified by different doses, types of clinical manifestation, the interaction between province and time from symptom onset to treatment initiation, and a representative center (**Figure 3**). Chloroquine showed beneficial effect in all stratum. However, the beneficial effect is not statistically significant in patients with severe COVID-19 symptoms, patients from Guangdong Province treated later than 14 days after symptom onset, or patients from SYSU5.

**Figure 3.**
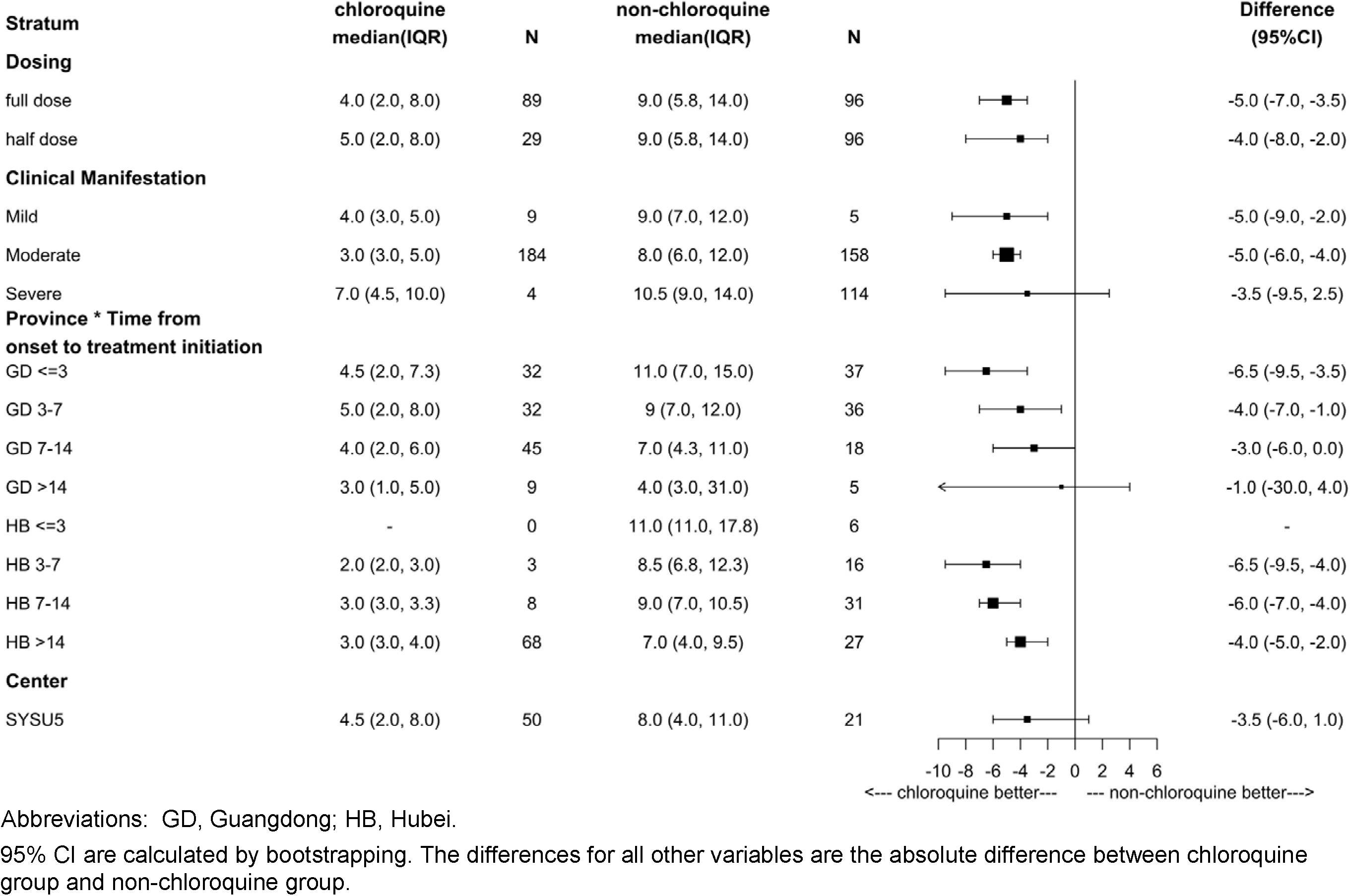
Post hoc analysis on the effect of chloroquine on time to undetectable viral RNA by stratification.

In order to assess the effect of chloroquine in more detailed clinical improvement outcomes in post hoc analysis, we collected detailed clinical data in patients from SYSU5, including the improvement of chest CT, the monitoring of serum chloroquine concentration, and the reappearance of positive viral RNA detection after hospital discharge. In this subgroup of patients, the interval time between symptom onset and treatment initiation were comparable. The medians are 7 days in chloroquine group (N=50) and 6 days in non-chloroquine group (N=21) (absolute difference in medians 1 day; 95% CI −3 to 4 days; *P* = 0.99; **Supplemental Table 3**). We did not find statistically significant difference in the time to undetectable viral RNA between the two groups (absolute difference in medians −3.5 days; 95% CI −6 to 1 days). The chloroquine group have higher percentage of patients with improved chest CT by day 10 (absolute difference in proportions 9.7; 95% CI −16.0 to 35.6) and day 14 (absolute difference in proportions 6.3; 95% CI −22.2 to 32.0) than the non-chloroquine group but the difference is not statistically significant (**Supplemental Table 3**). This could be due to the small sample size or the delayed chest CT absorption^9^. We did not observe beneficial effect of chloroquine in the length of hospital stay and the duration of oxygen support (**Supplemental Table 3**). Unprecedently, we observed 3 cases of so called “re-positive” patients in the chloroquine group. They were identified with negative viral RNA test from respiratory tract samples but positive viral RNA test from fecal samples within 7 days following hospital discharge. No such observation in the non-chloroquine group. Investigation is underway to examine whether it is due to re-infection or other factors.

Among the 12 hospitals, one hospital explored different dosage of chloroquine, as 500 mg once daily, which is half of the protocol dosage. We compared the primary and secondary outcomes in patients from this subgroup (N=29) with the non-chloroquine group in Guangdong Province. The results mainly showed that chloroquine has benefit effect on the time to undetectable viral RNA (absolute difference in medians −5 days; 95% CI −6.0 to −4.0 days) and the proportion of patients with undetectable viral RNA by day 10 is higher in chloroquine group (absolute difference in proportions 32.7; 95% CI 23.9 to 42.1). The duration of fever was also shorter than those in the non-chloroquine group (geometric mean ratio 0.8; 95% CI 0.5 to 0.9) (**Supplemental Table 4**).

### Safety

A total of 53 patients (26.9%) in the chloroquine group and 57 (32.4%) in the non-chloroquine group reported adverse events during study period (**Table 3**). Gastrointestinal events including vomiting, abdominal distension, nausea, decreased appetite, thirst were more common in chloroquine than in the non-chloroquine group. The percentage of patients with neurological adverse events, including dizziness and sleep order, were higher in the chloroquine than in the non-chloroquine group. In addition, anxiety was observed more frequently in chloroquine than in the non-chloroquine group. We observed fewer adverse events in patients with half dose of chloroquine than full dose (absolute difference in proportions −40; 95% CI −60 to −29).

**Table 3.**
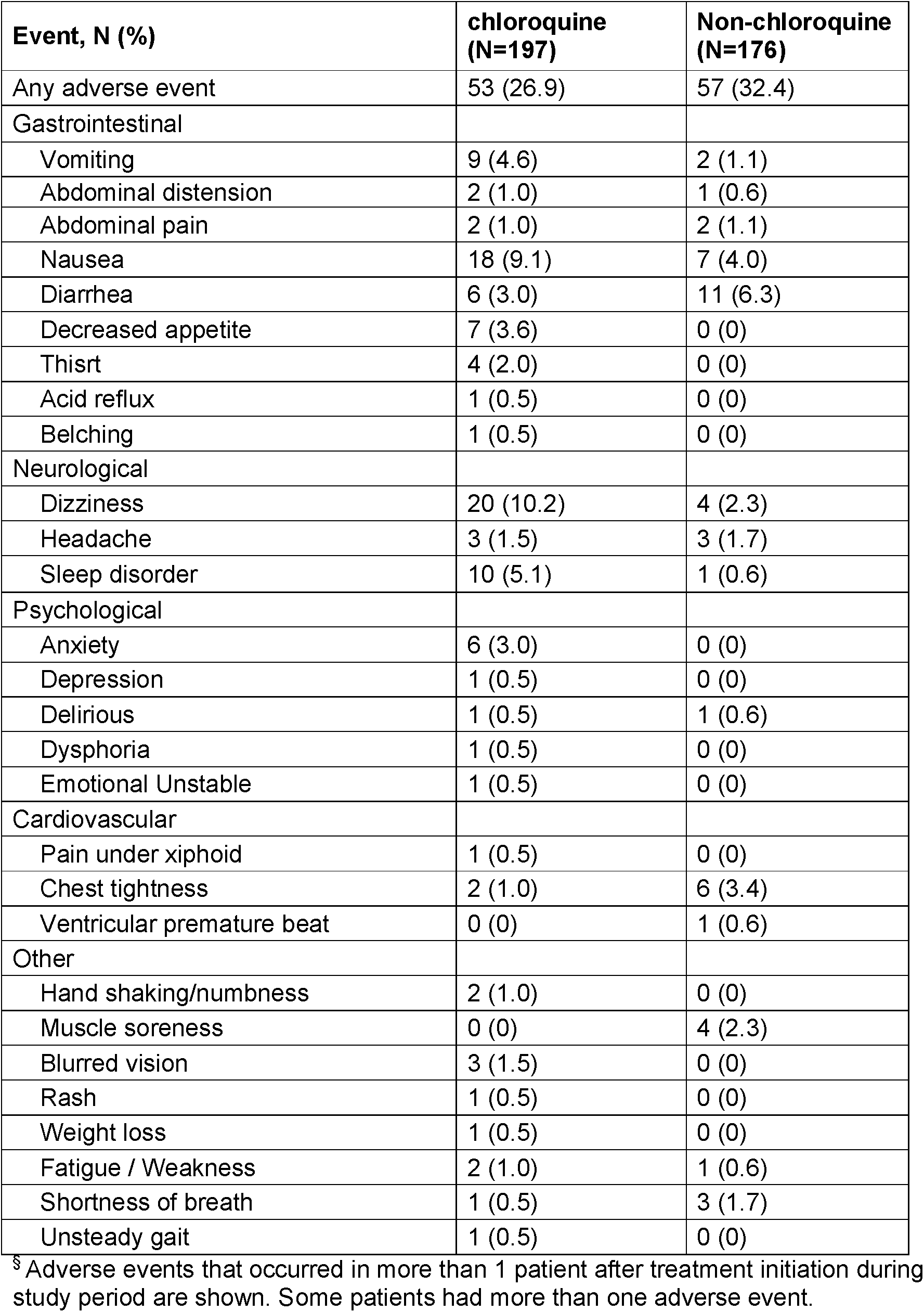
Summary of adverse events^§^.

| **Event, N (%)** | **chloroquine**<br>**(N=197)** | **Non-chloroquine**<br>**(N=176)** |
| --- | --- | --- |
| Any adverse event | 53 (26.9) | 57 (32.4) |
| Gastrointestinal |  |  |
| Vomiting | 9 (4.6) | 2 (1.1) |
| Abdominal distension | 2 (1.0) | 1 (0.6) |
| Abdominal pain | 2 (1.0) | 2 (1.1) |
| Nausea | 18 (9.1) | 7 (4.0) |
| Diarrhea | 6 (3.0) | 11 (6.3) |
| Decreased appetite | 7 (3.6) | 0 (0) |
| Thirst | 4 (2.0) | 0 (0) |
| Acid reflux | 1 (0.5) | 0 (0) |
| Belching | 1 (0.5) | 0 (0) |
| Neurological |  |  |
| Dizziness | 20 (10.2) | 4 (2.3) |
| Headache | 3 (1.5) | 3 (1.7) |
| Sleep disorder | 10 (5.1) | 1 (0.6) |
| Psychological |  |  |
| Anxiety | 6 (3.0) | 0 (0) |
| Depression | 1 (0.5) | 0 (0) |
| Delirious | 1 (0.5) | 1 (0.6) |
| Dysphoria | 1 (0.5) | 0 (0) |
| Emotional Unstable | 1 (0.5) | 0 (0) |
| Cardiovascular |  |  |
| Pain under xiphoid | 1 (0.5) | 0 (0) |
| Chest tightness | 2 (1.0) | 6 (3.4) |
| Ventricular premature beat | 0 (0) | 1 (0.6) |
| Other |  |  |
| Hand shaking/numbness | 2 (1.0) | 0 (0) |
| Muscle soreness | 0 (0) | 4 (2.3) |
| Blurred vision | 3 (1.5) | 0 (0) |
| Rash | 1 (0.5) | 0 (0) |
| Weight loss | 1 (0.5) | 0 (0) |
| Fatigue / Weakness | 2 (1.0) | 1 (0.6) |
| Shortness of breath | 1 (0.5) | 3 (1.7) |
| Unsteady gait | 1 (0.5) | 0 (0) |

<sup>§</sup> Adverse events that occurred in more than 1 patient after treatment initiation during study period are shown. Some patients had more than one adverse event.

Chloroquine phosphate has a long half-life (20-60 days)^10-12^ and its mean residence time is approximately 20 days^10^. It may have cumulative effect^13^. In order to determine whether chloroquine has a cumulative effect in the short-term treatment with COVID-19, we measured the serum concentration of chloroquine in patients from SYSU5 during and off the treatment. The results showed that the mean of serum concentration of chloroquine gradually rising, with the highest reaching 1.80(±0.49) μmol/L during medication and reduced to 0.13(±0.08) μmol/L within 28±1 days off chloroquine (**Supplemental Figure 3**).

## DISCUSSION

In this study, we found that patients in the chloroquine group experienced significantly faster and higher rate of viral suppression comparing to the non-chloroquine group in both the full analysis and the post hoc stratified analysis. Even when the dose reduced to half, the benefit of chloroquine still remained (**Figure 3**). These findings indicate that chloroquine could be effective in treating patients with COVID-19. To our knowledge, this is the first and largest clinical study on chloroquine phosphate for treating COVID-19 to date.

We recognize that our study has several limitations. This study was carried out under the COVID-19 public health emergency. Due to the limited medical capacity and urgent clinical situation, we were unable to conduct a standard randomised controlled study to formally evaluate efficacy and safety of chloroquine versus placebo. As an observational study, we have to note that several factors may influence the interpretation of the result. It is reasonable to suspect that the dramatic improvement in the primary outcome in chloroquine could be due to the later treatment initiation since symptom onset. Firstly, gaining experience in treatment management and attenuation of the virus during the course of the epidemic could contribute to the improved outcomes. Secondly, we cannot rule out the possibility that among those with longer interval time between symptom onset and treatment, some may already have been on the course of recovery. Nevertheless, post hoc analysis dividing subgroups according to the interval time did not change the conclusion that the chloroquine group had a better outcome than the non-chloroquine group. Notably, some of the strata were incomparable due to small sample size. Thirdly, although it is impossible to dissect the influence from other antiviral therapies used before chloroquine, it is a plausible assumption that chloroquine is the first antiviral therapy used in the group of patients treated within 3 days since symptom onset. In this stratum, chloroquine still benefits patients with faster viral suppression (**Figure 3**). Lastly, due to the differences in personnel and technical equipment of among all hospitals, we could not fully collect clinical and laboratory data of all patients. However, detailed clinical data were obtained from the chloroquine patients enrolled from SYSU5, enabling advanced analysis of clinical outcomes and pharmacokinetics.

As of this time, there are more than 20 trials ongoing for evaluating the efficacy and safety of chloroquine or hydroxychloroquine in treating COVID-19. Magagnoli et al. recently published a retrospective study indicating that the use of hydroxychloroquine with or without azithromycin does not reduced the risk of mechanical ventilation in United States veterans hospitalized with COVID-19^14^. Comparing with this study, our study population included both genders, was much younger, has fewer patients with severe symptoms that requires ventilation. Therefore, prospective randomised trials are needed to see if the results can be replicated.

Till now, the mechanism of chloroquine’s effect agianst SARS-CoV-2 remained unelucidated. Clatherin-mediated endocytosis is required for entry of coronavirus into host cells and meanwhile autophagy involves in viral replication^15^. Chloroquine inhibits clatherin-mediated endocytosis by suppressing acidification of endosomes, and autophagy by raising its lysosomal PH and blocking fusion of autophagosome with lysosome and lysosomal protein degradation^16^. A recent study has shown that the development of COVID-19 disturbed metabolic patterns, which aligned with the progress and severity of COVID-19 (Wu et al. National Science Review 2020, in press). Chloroquine has a favorable effect on glucose and lipid metabolism^17^. Therefore, chloroquine may exert its antiviral effect against SARS-CoV-2 by inhibiting endocytosis and autophagy, and stabilizing glucose and lipid metabolism.

The adverse reactions of chloroquine drugs are of great concern to the community. Although it is an old anti-malarial drug, its safety in treating COVID-19 patients is still unknown. In the present study, we did not observe serious adverse events in patients with chloroquine. All adverse events observed during the study period are known side-effects for chloroquine (**Table 3**). The main adverse events were symptoms in gastrointestinal and neuropsychiatric systems. Chloroquine is known for its side effects in cardiovascular system. In the chloroquine group, we did not find significantly higher rate of adverse events in patients older than 65 or with pre-existing conditions (**Supplement Table 5**). Adverse event appeared in 1 out of 29 patients (3.5%) with half dose while in 52 out 168 patients (31.0%) with full dose, indicating that the half dose group has lower adverse event rate (absolute rate difference −27.5; 95% CI −45.0 to −19.2). Although previous studies suggested that chloroquine may have cumulative effect^11,18,19^, we did not observe cumulative effects among 50 patients from SYSU5 by monitoring the serum concentration of chloroquine for up to 28 days after treatment completion. Future studies are needed to determine the optimal dosing for treating COVID-19 and the cumulative effect of chloroquine in tissues and organs. Severe cases are underrepresented in the present study, and thus should be focused in the future studies to evaluate the efficacy and safety profile in this population. In addition, it will be important to study the prophylaxical use of chloroquine in areas with high rate of COVID-19 or in health professionals working with COVID-19 patients.

In conclusion, our preliminary evidence showed that chloroquine has the potential to shorten the time to SARS-CoV-2 viral suppression and duration of fever, even with reduced dose. Further randomised studies are needed to determine the optimal dose, to assess its benefit for both severe cases and to assess its benefit in settings other than secondary care. Considering that there is no better option at present, chloroquine could be a viable option to combat the coronavirus pandemic under proper management.

## METHODS

### Study Design and participants

This study was a multicenter prospective observational study conducted from February 7 through March 8, 2020 at 11 hospitals in Guangdong Province and 1 mobile cabin hospital in Wuhan, Hubei Province, China. The study protocol was approved by the ethics committee of Fifth Affiliated Hospital of Sun Yat-sen University (SYSU5), located in Zhuhai, Guangdong Province, and registered at Chinese Clinical Trial Registry (ChiCTR2000029609). We did this study in accordance with the principles of the Declaration of Helsinki and Good Clinical Practice. Written informed consent was obtained from all patients or their legal guardians. During the study period, each hospital had various choices of antiviral regimen, and the sample size of Lopinavir/Ritonavir (the historical control group in the original protocol) for single-use were underpowered. Thus, we updated the inclusion criteria of the historical control group as patients receiving non-chloroquine treatment.

Eligible patients were aged 18 years or older with confirmed SARS-CoV-2 infection, tested by the local Center for Disease Control (CDC) or by a designated diagnostic laboratory, using reverse-transcriptase-polymerase-chain-reaction (RT-PCR) assay (Shanghai ZJ Bio-Tech Co Ltd) for SARS-CoV-2 in a respiratory tract sample. Patients were ineligible if he/she met any of the following criteria: pregnant women, with known allergies to 4-aminoquinoline compounds, blood system diseases, chronic liver or kidney diseases in end-stage, arrhythmia or second/third degree heart block, with known to have retinopathy, hypoacusis or hearing loss, mental disease, glucose-6-phosphate dehydrogenase (G6PD) deficiency, had received digitalis drugs within the 7 days preceding enrollment, or is classified as critical case according to China’s Novel Coronavirus Pneumonia Diagnosis and Treatment Plan (4^th^ Edition). Enrolled patients received 500mg chloroquine Phosphate (equivalent of 300 mg chloroquine base, Shanghai Xinyi Pharmaceutical Co., Ltd) orally, once/twice-daily with no other antiviral therapies. The criteria of stopping chloroquine was defined as undetectable viral RNA for two consecutive respiratory tract samples. The duration of medication in chloroquine group is no more than 10 days. Patients in the historical control group were treated according to China’s Novel Coronavirus Pneumonia Diagnosis and Treatment Plan (details described in **Supplemental Table 6**).

### Outcome and measurements

The primary outcome is the time from treatment initiation to undetectable viral RNA for two consecutive respiratory tract samples. The secondary outcomes include the proportion of patients with undetectable viral RNA by day 10 and 14, duration of fevers, time in hospital, and adverse events. The detailed definition of outcomes is described in **Supplementary Methods**. Respiratory tract sample was collected from patients daily to conduct RT-PCR assay for SARS-CoV-2 infection. The epidemiological characteristics, clinical symptoms and signs, adverse reactions/events were collected with data collection forms. The outcomes, clinical characteristics, laboratory findings, chest computed tomographic (CT) scans were recorded on case record forms and then double-entered into an electronic database and validated by trial staff. After hospital discharge, patients were followed up once weekly. Patients with “re-positive” viral RNA detection within one week after hospital discharge are defined as having either 2 consecutive RT-PCR positive result from either respiratory tract sample or fecal specimen. In the subgroup of patients in SYSU5, all CT images were reviewed by two fellowship-trained cardio-thoracic radiologists by using a viewing console. Images were reviewed independently, and final decisions were reached by consensus ^9^.

To fully assess the safety of chloroquine, we monitor the serum concentration of chloroquine at the day 1, 3, 5, 7, 10 during drug administration and day 1 to 7, and day 14, day 21 after treatment completion in a subgroup of samples enrolled from SYSU5 (N=50). Details about the measurement of serum concentration of chloroquine are described in **Supplemental Methods**.

### Statistical Analysis

The original plan was to compare the efficacy between three groups, chloroquine only, Lopinavir/Ritonavir only, and chloroquine plus Lopinavor/Ritonavir. At the beginning of the outbreak, different therapies were proposed and tested for the treatment of COVID-19. Therefore, it is challenging to find sufficient patients with unified treatment across all centers. The epidemic in Guangdong had been brought under control rapidly during the study making it difficult to recruit patients as planned. The history of changes to the protocol is listed in **Supplemental Table 7**. Thus, a decision was made to focus on recruiting chloroquine only and compare the efficacy with historical controls. The current sample size was based on feasibility within the fixed trial recruitment window and was felt would provide sufficient precision for the estimation of plausible effects. With right-censoring in time-to-event variables, generalized Wilcoxon test was used to compare the difference in medians and the 95% confidence intervals were calculated by bootstrapping^20^. For binary outcomes, Wilson test was implemented to calculate the difference in proportions and 95% confidence intervals. As this was an observational study, imbalance in the baseline characteristics of the two groups was expected. To adjust for this imbalance, we performed post hoc analyses within various subgroups by two dosage options, by clinical manifestation, by the interaction of province and the interval time between symptom onset and treatment initiation (≤ 3 days; 3~7 days; 7~14 days; > 14 days), and by center. For all comparative analyses, *P* <0.05 was considered statistically significant. No allowance for multiplicity. All P values are two tailed. All statistical analyses were performed in R, version 3.6.1 (R Foundation for Statistical Computing)^21^.

### Role of the funding source

The sponsor of the study had no role in study design, data collection, data analysis, data interpretation, or writing of the report. The corresponding author had full access to all the data and had final responsibility for the decision to submit for publication.

## Data Availability

The data that support the findings of this study are available from the corresponding author on reasonable request. Participant data without names and identifiers will be made available after approval from the corresponding author and Ministry of science and technology and Health Committee in Guangdong province. After publication of study findings, the data will be available for others to request. The research team will provide an email address for communication once the data are approved to be shared with others. The proposal with detailed description of study objectives and statistical analysis plan will be needed for evaluation of the reasonability to request for our data. The corresponding author and Ministry of science and technology and Health Committee in Guangdong province will make a decision based on these materials. Additional materials may also be required during the process.

## Contributors

S.H, N.Z, S.J. J.X. L.T and D.P. had the idea for and designed the study and had full access to all data in the study and take responsibility for the integrity of the data and the accuracy of the data analysis. M.L, F.X, Y.L., M.H, J.L, P.P and T.T contributed to writing of the report. M.L, F.X, M.H, Y.L., J.L and P.P contributed to critical revision of the report. M.L contributed to the statistical analysis. All authors contributed to data acquisition, data analysis, or data interpretation, and reviewed and approved the final version.

## Declaration of interest

All authors declare no competing interests.

## Acknowledgements

We thank all patients who participated in this study and their families. We also thank all the health care workers on the front lines of the COVID-19 pandemic. This work was supported in part by National key Research and Development (R&D plan “public security risk prevention and control and emergency technical equipment”: Novel coronavirus infection pneumonia diagnosis and control key technology research (2020YFC082400), Guangdong Research and Development (R&D) Program in Key Areas: Guangdong Scientific and Technological Research Special fund for Prevention and Treatment of COVID-19, Special fund for emergency research on SARS-CoV-2 from the Guangzhou Regenerative Medicine and Health Guangdong Laboratory, Cultivation Project of “Three Major Scientific Research Projects” of Sun Yat-sen University in 2020, Emergency Project of New Coronavirus Prevention and Control Science and Technology of Sun Yat-sen University, and Zhuhai 2020 “novel coronavirus infection control” emergency technology tackling key issues. We thank all the participants in this study. We acknowledge all health-care workers who provided care for the patients at the 12 hospitals. We thank Jiaxing Taimei Medical Technology Co., Ltd for Electronic Data Capture service.

## Notes

### Competing Interest Statement

The authors have declared no competing interest.

## References

1. Lu H, Stratton CW, Tang YW. Outbreak of pneumonia of unknown etiology in Wuhan, China: The mystery and the miracle. J Med Virol 2020; 92(4): 401–2.

2. Hui DS, E Ia, Madani TA, et al. The continuing 2019-nCoV epidemic threat of novel coronaviruses to global health - The latest 2019 novel coronavirus outbreak in Wuhan, China. Int J Infect Dis 2020; 91: 264–6.

3. Zhu N, Zhang D, Wang W, et al. A Novel Coronavirus from Patients with Pneumonia in China, 2019. N Engl J Med 2020; 382(8): 727–33.

4. Keyaerts E, Vijgen L, Maes P, Neyts J, Van Ranst M. In vitro inhibition of severe acute respiratory syndrome coronavirus by chloroquine. Biochem Biophys Res Commun 2004; 323(1): 264–8.

5. Kono M, Tatsumi K, Imai AM, Saito K, Kuriyama T, Shirasawa H. Inhibition of human coronavirus 229E infection in human epithelial lung cells (L132) by chloroquine: involvement of p38 MAPK and ERK. Antiviral Res 2008; 77(2): 150–2.

6. Wang M, Cao R, Zhang L, et al. Remdesivir and chloroquine effectively inhibit the recently emerged novel coronavirus (2019-nCoV) in vitro. Cell Res 2020; 30(3): 269–71.

7. Cortegiani A, Ingoglia G, Ippolito M, Giarratano A, Einav S. A systematic review on the efficacy and safety of chloroquine for the treatment of COVID-19. J Crit Care 2020.

8. Organization WH. Clinical management of severe acute respiratory infection when novel coronavirus (2019-nCoV) infection is suspected: interim guidance, 28 January 2020: World Health Organization, 2020.

9. Chung M, Bernheim A, Mei X, et al. CT Imaging Features of 2019 Novel Coronavirus (2019-nCoV). Radiology 2020; 295(1): 202–7.

10. Gustafsson L, Lindstrom B, Grahnen A, Alvan G. Chloroquine excretion following malaria prophylaxis. British Journal of Clinical Pharmacology 1987; 24(2): 221–4.

11. Krishna S, White NJ. Pharmacokinetics of quinine, chloroquine and amodiaquine. Clinical implications. Clin Pharmacokinet 1996; 30(4): 263–99.

12. Ducharme J, Farinotti R. Clinical pharmacokinetics and metabolism of chloroquine. Focus on recent advancements. Clin Pharmacokinet 1996; 31(4): 257–74.

13. Marks JS. Chloroquine retinopathy: is there a safe daily dose? Ann Rheum Dis 1982; 41(1): 52–8.

14. Magagnoli J, Narendran S, Pereira F, et al. Outcomes of hydroxychloroquine usage in United States veterans hospitalized with Covid-19. *medRxiv2020:* 2020.04.16.20065920.

15. Yang N, Shen HM. Targeting the Endocytic Pathway and Autophagy Process as a Novel Therapeutic Strategy in COVID-19. Int J Biol Sci 2020; 16(10): 1724–31.

16. Shintani T, Klionsky DJ. Autophagy in health and disease: a double-edged sword. Science 2004; 306(5698): 990–5.

17. Hage MP, Al-Badri MR, Azar ST. A favorable effect of hydroxychloroquine on glucose and lipid metabolism beyond its anti-inflammatory role. Ther Adv Endocrinol Metab 2014; 5(4): 77–85.

18. Augustijns P, Geusens P, Verbeke N. Chloroquine levels in blood during chronic treatment of patients with rheumatoid arthritis. European Journal of Clinical Pharmacology 1992; 42(4): 429–33.

19. Akpovwa H. Chloroquine could be used for the treatment of filoviral infections and other viral infections that emerge or emerged from viruses requiring an acidic pH for infectivity Cell Biochem Funct 2016; 34(4): 191–6.

20. Keene ON. Alternatives to the hazard ratio in summarizing efficacy in time-to-event studies: an example from influenza trials. Statistics in Medicine 2002; 21(23): 3687–700.

21. Team RC. R: A language and environment for statistical computing. 2013.

